# No Evidence of Anti-influenza Nucleoprotein Antibodies in Retail Milk from Across Canada (April – July 2024)

**DOI:** 10.1101/2025.01.31.25321461

**Authors:** Hannah L. Wallace, Jordan Wight, Gabriela J. Rzeszutek, Mustafa S. Jafri, Mariana Baz, Barbara Dowding, Louis Flamand, Tom Hobman, François Jean, Jeffrey B. Joy, Andrew S. Lang, Sonya MacParland, Craig McCormick, Ryan Noyce, Rodney S. Russell, Selena M. Sagan, Jumari Snyman, Isaac I. Bogoch, Angela L. Rasmussen, Anne W. Rimoin, Jason Kindrachuk, the Pan-Canadian Milk Study Network

## Abstract

Following reports of HPAI H5N1 infections of dairy cattle in the United States (US) in March 2024, we established a Pan-Canadian Milk network to monitor retail milk in Canada. Milk samples from across Canada that had previously tested negative for influenza A virus (IAV) RNA were tested for the presence of anti-IAV nucleoprotein (NP) antibodies, as an indicator of past infection of dairy cattle. None of the 109 milk samples tested had evidence of anti-IAV NP antibodies. This is consistent with previous findings from our academic group as well as others including federal testing initiatives that have not found any IAV RNA in milk. Although not surprising given that no cases of H5N1 in cattle have been reported in Canada to date, this work further supports that the extensive outbreak in dairy cattle in the US has not extended northward into Canada, and the integrity of the Canadian milk supply remains intact.

## 1.0 Introduction

Highly pathogenic avian influenza (HPAI) H5N1 (A/goose/Guangdong/1996) clade 2.3.4.4b was introduced into North America at the end of 2021, with initial cases detected in Newfoundland and Labrador, Canada in wild birds and at an exhibition farm in the region (Caliendo et al., 2022). Subsequently, the virus spread extensively throughout the Americas and to Antarctica, infected a broad diversity of species. This has resulted in numerous mass mortality events in multiple species and populations of wild birds as well as terrestrial and marine mammals (Alkie et al., 2023; European Food Safety Authority et al., 2024; Jakobek et al., 2023; Lair et al., 2024; Peacock et al., 2025; Plaza et al., 2024; Youk et al., 2023).

The species expansion of HPAI H5N1 has included an unprecedented outbreak of the virus in dairy cattle that began in Texas, United States (US) in March 2024 (Hu et al., 2024; Nguyen et al., 2024; USDA APHIS, 2025). The virus was found to be similar to those circulating in wild birds in the region at the time, indicating a likely spillover from wild birds into dairy cattle (Worobey et al., 2024). Since the initial cases in Texas, 16 states have reported outbreaks in cattle, with 929 herds reported to be infected as of 20 January 2025 (USDA APHIS, 2025). Spread of the virus has continued despite restrictions on movement of infected cattle between states (USDA APHIS, 2024). Due to the number of infected herds, the state of California declared a State of Emergency in December 2024 in response to half of the dairy cattle herds in the state reporting cases of H5N1 (California Department of Public Health, 2025).

While dairy cattle display relatively mild signs of H5N1 infection, including lethargy, mild respiratory symptoms, decreased feed intake, dehydration, decreased milk production, and production of milk with abnormal colour/texture (Baker et al., 2024; Burrough et al., 2024; Caserta et al., 2024), infections of other animals such as barn cats have been associated with high fatality rates (Burrough et al., 2024). High fatality rates (up to 50%) have also historically been associated with infections of humans with H5N1 viruses, however this has not been the case with the current outbreak of viruses associated with US cattle and poultry (World Health Organization, 2024). Since March 2024, there have been 66 human cases of H5N1 clade 2.3.4.4b reported in the US, primarily related to outbreaks in either dairy cattle or poultry facilities. While the majority of the 66 known individuals who have become infected have had only mild symptoms and recovered fully, there has been one human fatality reported who was known to have direct exposure to infected poultry (Centers for Disease Control and Prevention, 2025a, 2024; Uyeki et al., 2024). Importantly, no human-to-human transmission has been reported in the US to date (Centers for Disease Control and Prevention, 2025b).

Although the current outbreak in the US was the first time extensive infections and spread of H5N1 in dairy cattle had been reported, infections of cattle with influenza A viruses (IAVs) have been reported since the 1950s (Brown et al., 1998; Campbell et al., 1977; Kalthoff et al., 2008; Mitchell et al., 1953). Indeed, even decreased milk production had been previously associated with IAV infection and rising anti-IAV antibody levels in cattle (Crawshaw et al., 2008). Although commonly thought of as being found in serum, antibodies can also be detected in milk secreted by lactating mammals. This enables passive exchange of protective antibodies from mother to immune naïve offspring, including in dairy cattle and their milk used for human consumption (Nobrega et al., 2023; Ploegaert et al., 2011). Therefore, presence of antibodies in blood or milk against specific pathogens can provide evidence of past infection.

Despite the geographic proximity of the US and Canada, to date, there have been no reported infections of H5N1 in dairy cattle in Canada. Our group, as well as an academic group in Ontario, and the Canadian Food Inspection Agency (CFIA), began testing retail and unpasteurized milk from across the country for presence of IAV RNA. While our group as well as Blais-Savoie et al. both reported no detections of IAV RNA (matrix gene) in retail milk, the CFIA has reported no detections of HPAI in retail or unpasteurized milk (Blais-Savoie et al., 2024; Canadian Food Inspection Agency, 2024; Wallace et al., 2025). Given this, as well as the fact that birds are only transiently IAV RNA positive after infection (Wight et al., 2024), and it is currently unknown how long dairy cattle remain RNA positive following infection, other testing methods such as detection of antibodies may be more useful for documenting past infection with IAVs. Having collected retail milk samples from across Canada as part of our previous work (Wallace et al., 2025), we sought to test these samples for anti-IAV antibodies, as a more durable indicator of past infection of Canadian dairy cattle.

## 2.0 Methods

### 2.1 Screening for Anti-IAV NP Antibodies

109 retail milk samples from our previous study (Wallace et al., 2025) were screened for presence of anti-influenza antibodies using the IDEXX AI MultiS-Screen Ab test (IDEXX Canada, Product # 99-12119) blocking enzyme-linked immunosorbent assay (ELISA) which detects antibodies against the IAV nucleoprotein (NP) (Brown et al., 2009). The assay was performed as per manufacturer’s instructions, except that the milk samples were not diluted. A sample to negative control ratio (S/N) of < 0.5 was considered positive for anti-IAV NP antibodies.

### 2.2 Additional Controls

In addition to the positive control included with the kit, serum from a known anti-influenza NP positive American black duck (*Anas rubripes*) from St. John’s, Newfoundland and Labrador was also used (Wight et al., 2024), supplied by Dr. Andrew Lang (Memorial University). This serum was used to make a “spike-in” positive control (1:10 dilution of serum in milk) to ensure assay sensitivity was not inhibited by the protein and lipid rich milk.

### 2.3 Statistical Analyses

Statistical analysis was conducted using R v4.1.0 (R Core Team, 2021), and confidence intervals were determined using base R and confint functions.

## 3.0 Results

All 109 milk samples that tested negative for IAV RNA previously (Wallace et al., 2025), were also negative for anti-influenza NP antibodies (mean S/N ratio 1.11, 95% confidence interval 1.09-1.13, range 0.93-1.43) (**Table 1, Supplementary Data 1**). Milk samples were unlikely to contain even low levels of anti-IAV NP antibodies as indicated by the mean absorbance for the milk samples (1.14, 95% confidence interval 1.12-1.16) being further from the cut-off value for positivity than the negative control (1.05, 95% confidence interval 0.92-1.17). For the “spike-in” positive control with known positive serum, the mean S/N ratio for the serum diluted 1:10 with diluent was 0.26, and the S/N ratio for the serum diluted 1:10 with milk was 0.18, indicating the milk did not inhibit the assay but rather seemed to improve its sensitivity.

**Table 1.**
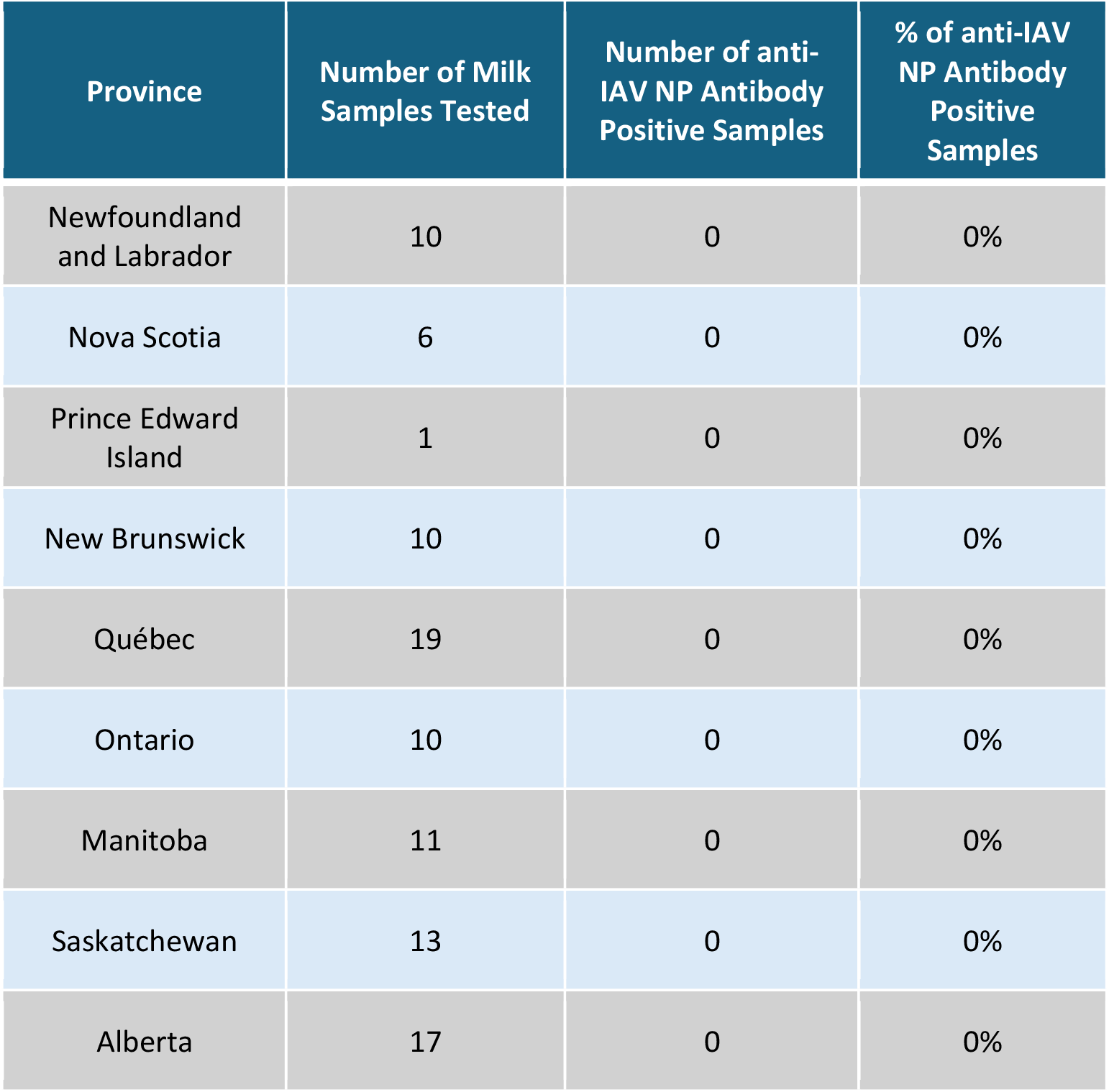
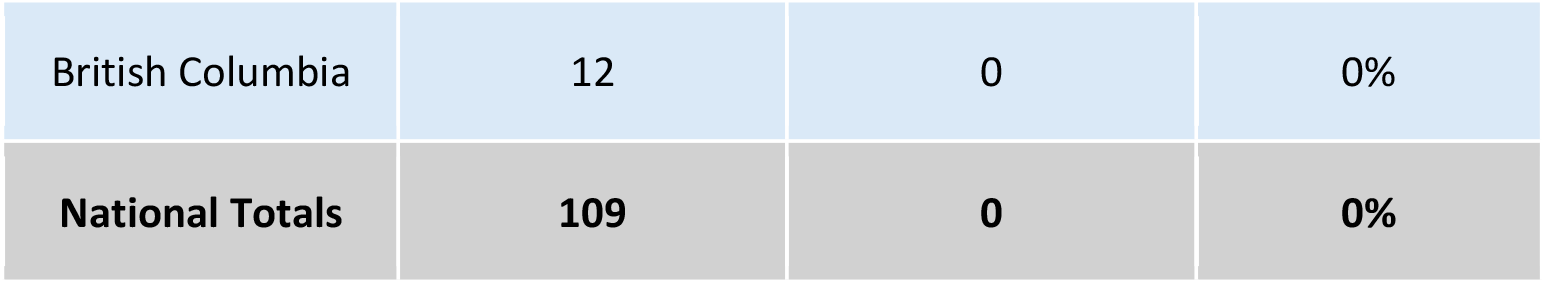
Provincial and national totals of retail milk samples tested, the number of samples positive for anti-IAV NP antibodies, and the percentage of anti-IAV NP antibody positive samples.

| Province | Number of Milk Samples Tested | Number of anti-IAV NP Antibody Positive Samples | % of anti-IAV NP Antibody Positive Samples |
| --- | --- | --- | --- |
| Newfoundland and Labrador | 10 | 0 | 0% |
| Nova Scotia | 6 | 0 | 0% |
| Prince Edward Island | 1 | 0 | 0% |
| New Brunswick | 10 | 0 | 0% |
| Québec | 19 | 0 | 0% |
| Ontario | 10 | 0 | 0% |
| Manitoba | 11 | 0 | 0% |
| Saskatchewan | 13 | 0 | 0% |
| Alberta | 17 | 0 | 0% |
| British Columbia | 12 | 0 | 0% |
| **National Totals** | **109** | **0** | **0%** |

## 4.0 Discussion

Lack of anti-IAV NP antibodies in retail milk tested in this study further support that IAVs are unlikely to have entered the Canadian milk supply via infected dairy cattle. This is consistent with data from our previous report (Wallace et al., 2025), as well as those of others (Blais-Savoie et al., 2024; Canadian Food Inspection Agency, 2024), that have not detected IAV RNA in milk in Canada.

This study employed a blocking ELISA to detect anti-IAV NP antibodies, and while it was originally developed and is validated for commercial poultry (IDEXX Laboratories, Inc., 2012), it has been used successfully to detect antibodies in a wide variety of species and used extensively for testing sera from wild and domestic animals (Brown et al., 2024; Damdinjav et al., 2025; Huang et al., 2014; Prosser et al., 2022; Stallknecht et al., 2024; Wight et al., 2024). As this kit has not been validated for use with milk, using it for this purpose may have affected the results, however this is unlikely given that presence of milk in our “spike-in” control did not negatively affect assay sensitivity. Additionally, while unknown how well antibodies present in cow milk would bind to the coated antigen of the plate, low level antibodies or those of low affinity would be expected to result in the S/N ratios differing much more greatly from the negative control, if they had been present.

As the number of H5N1-infected dairy herds continues to expand in the US, important questions persist about mechanisms of virus transmission and protective immune responses. Validation of the IDEXX assay for other species and sample types as well as development of new antibody detection assays, including against specific subtypes of IAVs, would be of significant value to improve our understanding of the extent of past infections in a broad diversity of potentially infected hosts. This is especially pertinent as the number of species that are reported to be susceptible to H5N1 continues to increase (Alkie et al., 2023; European Food Safety Authority et al., 2024; Jakobek et al., 2023; Lair et al., 2024; Peacock et al., 2025; Plaza et al., 2024; Youk et al., 2023). The lack of knowledge of past H5N1 (and other avian IAVs) infections in wild birds, mammals, and humans has been identified as one of the largest gaps in our collective knowledge about avian-origin IAVs (El Masry, I. et al., 2024).

## 5.0 Conclusions

Even though extensive transmission of H5N1 has occurred in dairy cattle herds in the neighbouring US, there continues to be no evidence of H5N1 infection of dairy cattle in Canada, with this study serving as the first investigation of the presence of anti-IAV antibodies in retail milk in Canada. Given ongoing transmission of H5N1 both among dairy cattle in the US and among wild birds across the globe there is ongoing risk of H5N1 being introduced into dairy cattle in Canada highlighting the need for continued vigilance and surveillance. Our pan-Canadian network of academic infectious disease researchers continues to be actively following the current situation and are ready to rapidly respond should H5N1 be detected directly in cattle and/or in the retail milk supply in Canada.

## Acknowledgements

We would like to acknowledge all PCM Study collaborators for contributing milk samples from across the country. We would also like to acknowledge the individuals who were involved in shipping milk samples, including Ishraq Rahman (Memorial University), Isabelle Dubuc (Université Laval), Carolina Camargo (University of British Columbia), Kim Pageau (Université Laval), Danielle G. Gordon (University of British Columbia), Ivan Villanueva (University of British Columbia), and Karla Fisher (Toronto General Hospital).

## Author Contributions

Conceptualization: HLW, JW, JK

Data Curation: HLW, JW

Formal Analysis: HLW, JW

Funding Acquisition: JK

Investigation: HLW, JW

Methodology: JW, HLW

Project Administration: HLW, JW, JK

Resources: JK, ASL Supervision: JK

Writing (original draft): HLW, JW

Writing (review and editing): All authors

## Data Availability

The raw data files will be made available from the authors upon request. S/N ratio data can be found in **Supplementary Data 1**.

